# Exposure to glucagon-like peptide 1 receptor (GLP-1R) agonists reduces glaucoma risk

**DOI:** 10.1101/2021.01.16.21249949

**Authors:** Jacob K. Sterling, Peiying Hua, Joshua L. Dunaief, Qi N. Cui, Brian L. VanderBeek

## Abstract

**Importance:** Glucagon-like peptide-1 receptor (GLP-1R) agonists regulate blood glucose and are commonly used to treat Type II Diabetes Mellitus. Recent work has shown that treatment with the novel GLP-1R agonist, NLY01, decreased retinal neuroinflammation and glial activation to rescue retinal ganglion cells in an animal model of glaucoma.

**Objective:** In this study, we used an insurance claims database to examine whether GLP-1R agonist exposure impacts glaucoma risk.

**Design, Setting, and Participants:** A retrospective cohort of adult patients who initiated a new GLP-1R agonist (i.e., exenatide, liraglutide, albiglutide, dulaglutide, semaglutide, or lixisenatide) was 1:3 age, gender, race, active diabetes medication classes, and year of index date matched to a cohort of patients who initiated a different class of oral diabetic medication during their time in the database. Exclusion occurred for <2 years in the database, age <18 years old, no visit to an eyecare provider prior to the index date, a prior diagnosis of glaucoma, glaucoma suspect, or ocular hypertension, or prior glaucoma medication, procedure, or surgery. Diabetes severity was assessed using hemoglobin A1c and the Diabetes Complications Severity Index (DCSI), a validated metric based on six categories of diabetic complications. Inverse probability of treatment weighting (IPTW) was used within a multivariable Cox proportional hazard regression model to test the association between GLP-1R agonist exposure and the primary outcome. IPTW was derived from a propensity score model based on the DCSI, HbA1c, demographic factors and other systemic health conditions.

**Exposure:** Glucagon-like peptide 1 receptor agonist.

**Main Outcomes and Measures:** New diagnosis of primary open angle glaucoma, glaucoma suspect, or low tension glaucoma.

**Results:** Cohorts were comprised of 1,961 new users of GLP-1R agonists matched to 4,371 unexposed controls. After IPTW, age was the only covariate imbalanced (SMD >0.1) between cohorts. Ten new diagnoses of glaucoma (0.51%) were present in the GLP-1R agonist cohort compared to 58 (1.33%) in the unexposed controls. After adjustment, GLP-1R exposure conferred a reduced hazard of 0.54 (95%CI: 0.35-0.85, P =0.007), suggesting that GLP-1R agonists reduce the risk for glaucoma.

**Conclusions and Relevance:** GLP-1R agonist use was associated with a statistically significant hazard reduction for a new glaucoma diagnosis. Our findings support further investigations into the use of GLP-1R agonists in glaucoma prevention.

## Introduction

Glaucoma is the leading cause of irreversible blindness globally. Primary open angle glaucoma (POAG) is the most prevalent form of glaucoma and is projected to affect approximately 80 million people worldwide by 2040.^1^In about 60% of cases, POAG is associated with ocular hypertension.^2^However, the fraction of POAG associated with elevated IOP varies significantly across different ethnic and racial groups.^3^All available therapies for glaucoma target intraocular pressure (IOP) reduction through either medical or surgical means. However, disease progression can continue despite IOP normalization, and aggressive IOP lowering is associated with vision-threatening complications.^4,5^New therapies, targeting mechanisms of glaucoma beyond IOP reduction, are urgently needed to prevent permanent vision loss in patients who have exhausted existing treatment options.^2^>

Glucagon-like peptide 1 (GLP-1) is an incretin hormone that regulates blood glucose, weight, and satiety. Agents that increase GLP-1 receptor (GLP-1R) signaling, including GLP-1 analogs and dipeptidyl peptidase 4 (DPP-IV) inhibitors, have been developed for the treatment of type 2 diabetes mellitus (DM). GLP-1R agonists first gained FDA-approval in 2005 and include exenatide, liraglutide, albiglutide, dulaglutide, semaglutide, and lixisenatide. In addition to their effects in the periphery, GLP-1R agonists cross the blood-brain barrier and exert effects in the central nervous system, influencing feeding behavior, cellular proliferation, mitochondrial function, and neuroinflammation.^6^GLP-1R agonists are neuroprotective in mouse models of neurodegenerative diseases including Parkinson’s^7^and Alzheimer’s disease^8^. A randomized control trial of 45 patients with Parkinson’s disease found a statistically significant improvement in both motor and non-motor deficits among subjects treated with exenatide.^9^In a randomized control trial targeting Alzheimer’s disease, liraglutide improved cognitive function.^10^An additional trial testing GLP-1R agonists in Parkinson’s disease (ClinicalTrials.gov, identifier NCT04154072) is ongoing.

Recent work has identified shared mechanisms of neurodegeneration in animal models of Parkinson’s disease and glaucoma. In both diseases, neuroinflammation driven by glial cell activation contributes to neuron death.^7,11,12^Treatment with a novel GLP-1R agonist ameliorated neuroinflammation in animal models of both diseases to either reduce dopaminergic neuron death^7^or rescue retinal ganglion cells (RGC)^11^. In this study, we used an insurance claims database to examine whether GLP-1R agonist use impacted the risk for a new glaucoma diagnosis.

## Methods

### Data Source

Optum’s de-identified Clinformatics™ Data Mart Database was used for this study. This database contains the medical claims from commercial and Medicare Advantage insurance plans obtained from a large US insurer. All outpatient medical claims (inclusive of office visits, associated diagnoses, and laboratory testing) and demographic data for each beneficiary during their enrollment was accessible. The subset of data available for this study included all patients in the database from January 1, 2008 to June 30, 2019. The University of Pennsylvania Institutional Review Board declared this study exempt because it involves anonymized data with removal of protected health information.

### Cohorts

All patients over 18 years old who initiated a new GLP-1R agonist (i.e., exenatide, liraglutide, albiglutide, dulaglutide, semaglutide, and lixisenatide) were eligible for inclusion in the GLP-1R cohort. The date of the initial prescription fill was assigned as the index date of each patient. Exclusion occurred for those with less than two years in the plan prior to the index date or did not see an eye care provider at least once prior to the index date. Patients were also excluded for any history of a glaucoma, glaucoma suspect or ocular hypertension diagnosis, having a glaucoma procedure/surgery, or having an active prescription for a glaucoma medication in the 6 months prior to the index date. See **Supplemental Table 1** for all codes used in this study.

An unexposed comparison cohort was created from patients who initiated a new oral diabetic medication during their time in the plan. GLP-1R agonist patients were matched 3:1 on age, gender, race, and year of index date of the unexposed. To further equalize the cohorts, the unexposed cohort was also matched on the number of active diabetic medications at the time of the index date. An active medication was defined as the number of active prescriptions within 45 days of the index date (i.e., if a patient initiating a GLP-1R agonist medication was already using 2 other diabetic medications, then the matched unexposed patient was required to be initiating their 3rd (non-GLP-1R agonist) class of diabetic medication). Matched controls were also required to meet all inclusion and exclusion criteria outlined above.

### Outcomes and Covariates

The primary outcome of interest was an ICD-9 or 10 incident diagnosis code for primary open angle glaucoma, glaucoma suspect, or low tension glaucoma at any time after the index date. Covariates assessed at the time of the index date were age, sex, race, geographic location, yearly income, and education level. Systemic health states were created for hypertension, hypercholesterolemia, kidney disease (none/chronic/end stage renal disease), and the diabetes complications severity index (DCSI). The DCSI is a validated metric that is calculated using diagnosis codes from six categories of diabetic complications.^13^Lab values were also assessed for hemoglobin A1c level. Health care utilization in the year prior to the index date was used as a proxy of overall health status and was accounted for by counting every distinct day with at least one claim for an office visit. While smoking is not often directly coded in administrative datasets, we used smoking diagnosis codes, use of antismoking drugs, and Current Procedural Terminology (CPT) codes for smoking cessation counseling. This method has previously found smoking rates in administrative databases to be 10-11%, similar to that of the general population.^14^To account for differences in these baseline covariates between the exposed and unexposed cohorts, inverse probability of treatment weighting (IPTW) using the estimated propensity scores from all covariates was performed in all subsequent analyses.

### Statistical analysis

Descriptive statistics were used to analyze the data, with continuous variables reported using mean and standard deviation and categorical variables using frequency and percentage. A multivariable Cox proportional hazard regression was performed to determine the association between hazard of developing glaucoma and GLP-1R agonist exposure. Patients were censored if any of the above exclusion criteria occurred after the index date: initiation of the comparison cohort of medication, a gap of > 60 days or more in active prescription for the cohort medication, a new diagnosis for a different form of glaucoma not included as the outcome, the patient exited from the insurance plan, or the end of observation was reached. All data analyses were performed using SAS software (version 9.4; SAS Institute Inc, Cary NC).

## Results

After inclusion and exclusion criteria were met, 1,961 new users of GLP-1R agonists (**Figure 1**) were matched to 4,371 unexposed controls. After matching and inverse proportional treatment weighting (IPTW), covariates including race, gender, education, income, geographic location, diagnosis of hypertension, diagnosis of hypercholesterolemia, presence and severity of kidney disease, history of smoking, mean values of hemoglobin A1c, DSCI, and median days of healthcare utilization were balanced and comparable between cohorts (standard mean deviation [SMD] <0.1 for all comparisons; see **Table 1** for baseline and weighted characteristics of the study population). Age was the only the imbalanced covariat (SMD =0.109), with the mean weighted age of the unexposed cohort (56.1 ±12.72 years [Mean ±SD]) being slightly older than that of the exposed cohort (54.3 ±19.2 years; see **Table 1** for all baseline covariates). After IPTW, the unexposed cohort was 51.4% female, 78.2% White, 12.6% Black, 8.0% Hispanic, had 45.9% of patients with diabetic retinopathy, and a mean hemoglobin A1c of 7.92 (SD ±2.36). This compared to the exposed group after IPTW at 55.0% female, 78.7% White, 11.3% black, 8.2% Hispanic, had 45.7% of patients with diabetic retinopathy, and a mean hemoglobin A1c of 7.91 (±3.08).

**Table 1.** Baseline characteristics of each group by GLP1-agonist status.

| Variable | Non-user (n=4371) |  | User (n=1961) |  | Standardized mean difference |
| --- | --- | --- | --- | --- | --- |
|  | Unweighted | Weighted | Unweighted | Weighted |  |
| **Race** |  |  |  |  | 0.047 |
| Asian | 69 (1.58%) | 1.55% | 33 (1.68%) | 1.72% |  |
| Black | 542 (12.40%) | 12.61% | 234 (11.93%) | 11.31% |  |
| Hispanic | 330 (7.55%) | 7.57% | 157 (8.01%) | 8.23% |  |
| White | 3430 (78.47%) | 78.27% | 1537 (78.38%) | 78.74% |  |
| **Gender (female)** | 2271 (51.96%) | 51.41% | 1028 (52.42%) | 55.04% | 0.073 |
| **Age** | 55.63 (10.59) | 56.09 (12.72) | 55.43 (10.43) | 54.31 (19.19) | -0.109 |
| **Education** |  |  |  |  | 0.016 |
| Less than 12th Grade | 14 (0.32%) | 0.30% | 5 (0.25%) | 0.30% |  |
| High School Diploma | 1321 (30.22%) | 30.22% | 589 (30.04%) | 30.34% |  |
| Less than bachelor's degree | 2380 (54.45%) | 54.56% | 1079 (55.02%) | 54.18% |  |
| Bachelor's degree Plus | 638 (14.60%) | 14.61% | 286 (14.58%) | 14.93% |  |
| Unknown | 18 (0.41%) | 0.32% | 2 (0.10%) | 0.25% |  |
| **Income** |  |  |  |  | 0.016 |
| Unknown | 533 (12.19%) | 12.28% | 246 (12.54%) | 12.26% |  |
| <\$40K | 661 (15.12%) | 15.23% | 296 (15.09%) | 15.62% | |
| \$40K-\$49K | 260 (5.95%) | 5.83% | 108 (5.51%) | 5.98% | |
| \$50K-\$59K | 336 (7.69%) | 7.38% | 132 (6.73%) | 7.11% | |
| \$60K-\$74K | 481 (11.00%) | 10.52% | 189 (9.64%) | 10.48% | |
| \$75K-\$99K | 708 (16.20%) | 16.18% | 322 (16.42%) | 16.06% | |
| \$100K+ | 1392 (31.85%) | 32.59% | 668 (34.06%) | 32.49% | |
| **Geographic Division** |  |  |  |  | 0.039 |
| Mountain | 326 (7.46%) | 7.51% | 148 (7.55%) | 7.77% |  |
| Northeast | 314 (7.18%) | 6.75% | 113 (5.76%) | 6.72% |  |
| Pacific | 254 (5.81%) | 5.41% | 88 (4.49%) | 5.62% |  |
| South Atlantic | 1491 (34.11%) | 34.23% | 677 (34.52%) | 33.98% |  |
| Southern Midwest | 695 (15.90%) | 17.54% | 405 (20.65%) | 17.70% |  |
| Unknown | 4 (0.09%) | 0.06% | 0 (0.00%) | 0.00% |  |
| Upper Midwest | 1287 (29.44%) | 28.50% | 530 (27.03%) | 28.21% |  |
| **Diabetic retinopathy** | 1817 (41.57%) | 45.87% | 1094 (55.79%) | 45.74% | -0.003 |
| **Hypertension** | 3507 (80.23%) | 82.61% | 1722 (87.81%) | 82.23% | -0.010 |
| **Hypercholesterolemia** | 3735 (85.45%) | 87.29% | 1792 (91.38%) | 87.05% | -0.007 |
| **Kidney Disease** |  |  |  |  | 0.008 |
| No | 3105 (71.04%) | 66.49% | 1104 (56.30%) | 66.82% |  |
| CKD | 1188 (27.18%) | 31.30% | 798 (40.69%) | 30.94% |  |
| ESRD | 78 (1.78%) | 2.21% | 59 (3.01%) | 2.24% |  |
| **Smoking** | 1061 (24.27%) | 24.90% | 510 (26.01%) | 24.94% | 0.001 |
| **HbA1c level** (mean(SD)) | 7.82 (1.90) | 7.92 (2.36) | 8.12 (1.80) | 7.91 (3.08) | -0.006 |
| **# of days of health care usage** (mean(SD)) | 7.39 (6.26) | 8.11 (8.64) | 9.28 (6.70) | 8.22 (10.50) | 0.012 |
| **DSCI** (Mean(SD)) | 1.66 (1.98) | 1.81 (2.51) | 2.12 (2.16) | 1.83 (3.62) | 0.004 |
| **# of active DM med classes** (mean(SD)) | 1.66 (0.74) |  | 1.89 (0.82) |  |  |
| **Time prior to censoring/events** |  |  |  |  |  |
| Mean (SD) | 266.5 (299.8) |  | 143.9 (195.1) |  |  |
| Median (Q1-Q3) | 151 (91-326) |  | 84 (28-168) |  |  |
| **Number of events (new diagnosis of glaucoma or glaucoma suspect)** |  |  |  |  |  |
| No event | 4313 (98.67%) | 98.65% | 1951 (99.49%) | 99.60% |  |
| Had an event | 58 (1.33%) | 1.35% | 10 (0.51%) | 0.40% |  |

**Figure 1.**
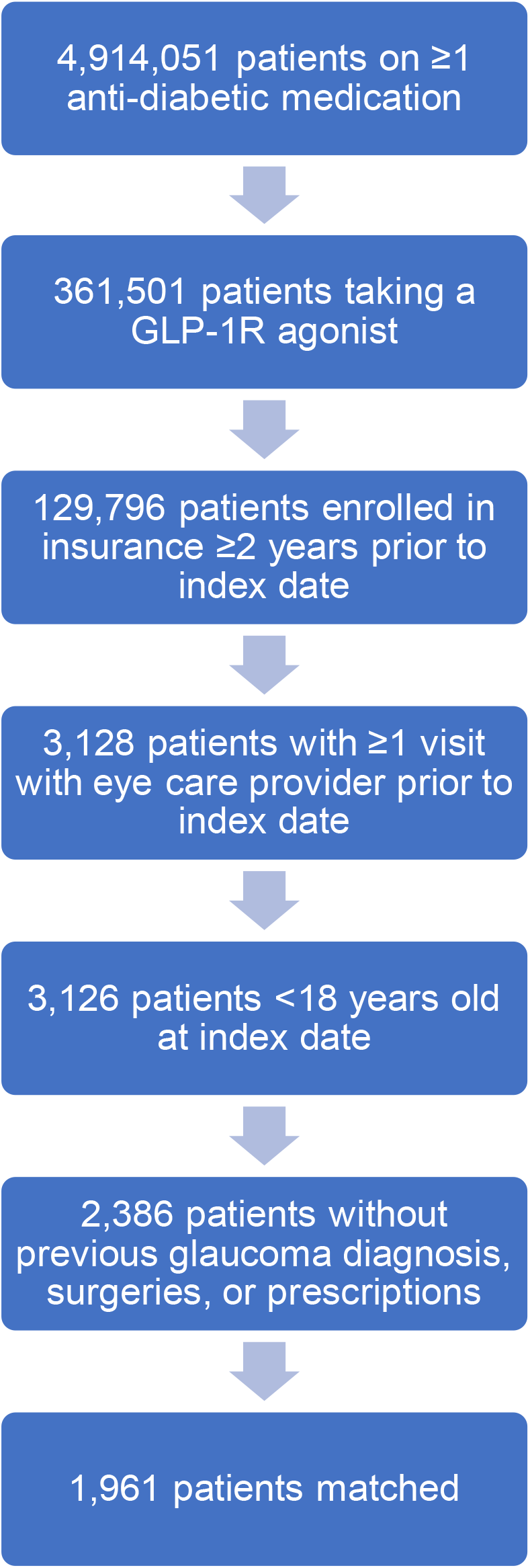
Flowchart showing inclusion/exclusion criteria for study patients.

During the follow-up period, 58 new diagnoses (1.33%) of glaucoma were present in unexposed controls compared to 10 new diagnoses (0.51%) of glaucoma in the GLP-1R agonist cohort. After IPTW of all covariates and the addition of age to the final multivariable model, Cox regression analysis revealed a 0.54 hazard ratio (95% CI: 0.35-0.85, P =0.007) for incident glaucoma among GLP-1R agonist patients versus unexposed controls. As expected, each year of increasing age was also associated with incident glaucoma (HR =1.03; 95%CI: 1.01-1.05, P <0.001; **Table 2**).

**Table 2.** Multivariable Cox regression analysis with inverse probability of treatment weighting.

| Variable | Category | Hazard Ratio (95%CI) | P-value |
| --- | --- | --- | --- |
| GLP-1R agonist |  |  | 0.007 |
|  | User | 0.54 (0.35, 0.85) |  |
|  | Non-user | Ref. |  |
| Age |  | 1.03 (1.01, 1.05) | <0.001 |

## Discussion

GLP-1R agonists are a common second-line therapy used in the treatment of type II diabetes mellitus (DM). In a mouse model of ocular hypertension, GLP-1R agonists can reduce RGC loss.^11^Using a national database, we examined the association between exposure to GLP-1R agonists and a new diagnosis of glaucoma or glaucoma suspect. Our analysis identified approximately 6,400 patients who fulfilled our inclusion criteria. Cox regression analysis showed a significant reduction in hazard for a new diagnosis of incident glaucoma in patients exposed to a GLP-1R agonist relative to unexposed controls. Result suggests that among DM patients, GLP-1R agonists may reduce the risk for glaucoma.

Preclinical data from a mouse model of ocular hypertension have shown that NLY01, a novel GLP-1R agonist designed to maximize CNS penetration, can reduce retinal inflammation and RGC death. Although the mechanism of protection has not been definitively determined, evidence suggests that NLY01 acts on microglia, macrophages, and perhaps astrocytes to reduce local retinal inflammation and prevent activation of the complement cascade after IOP elevation. Specifically, GLP-1R agonism reduces production of pro-inflammatory signaling molecules by microglia/macrophages, retinal levels of complement component 3 (C3), and reactive astrocyte formation.^11^Together these immunomodulatory effects may reduce RGC death in ocular hypertension.

Preclinical and randomized clinical trial data suggest that GLP-1R agonists may also be protective in neurodegenerative diseases of the brain, including Alzheimer’s^8^and Parkinson’s^7^disease. Although data on the neuroprotective role of GLP-1R agonists are relatively new, this class of medication is backed by more than 15 years of safety data due to their widespread use as DM therapy, making them an attractive therapeutic option for patients suffering from slowly progressive neurodegenerative diseases who may require decades of therapy to preserve neurological function of the retina and/or brain.

Our study focused on DM patients because GLP-1R agonists are FDA-approved to treat type II DM. However, it should be noted that multiple studies have shown an association between DM and an increased risk of POAG.^15–17^Despite this association, the mechanistic link between DM and glaucoma is not well understood and warrants additional analysis. Of note, our cohorts were comparable with respect to diabetes severity as evidenced by the diabetes complications severity index and hemoglobin A1c levels, reducing the potential confounding effect of DM in this study.

In addition to DM, increasing age is a risk factor for glaucoma.^4^In our study, there was a statistically significant difference in adjusted age between the two cohorts (Table 2). However, the difference in mean age was small (< 2 years), and unlikely to account for the observed difference in glaucoma hazard. In addition, the Cox proportional hazards model used in this study included terms derived from the weighted propensity scores, GLP-1R agonist exposure and age.

Although our results show a statistically significant reduction in hazard for a new glaucoma or glaucoma suspect diagnosis among patients treated with GLP-1R agonists, our analysis identified only 68 patients across both the unexposed and exposed cohorts who met outcome criteria. This was largely due to censoring for patients with a gap in active prescription longer than 60 days. Although this requirement limited the number of patients who met outcome criteria, to alter this requirement would mean including more patients who were not actively taking a GLP-1R agonist and would bias the study towards the null (i.e., exposed and unexposed becoming more similar due to lack of exposure). Despite this limitation, we still found a reduction in hazard in those treated with GLP-1R agonists, attesting to the robustness of this association.

Further, lack of specific clinical data including visual acuity prevented us from examining whether GLP-1R agonists reduced the risk of glaucoma progression or improved visual outcomes. Future work utilizing a larger database containing ocular data, including measures of glaucoma severity, would address these limitations and provide a more complete picture of the potential for GLP-1R agonist-mediated neuroprotection in glaucoma.

Our findings, in combination with preclinical data, support further investigations into the use of GLP-1R agonists for glaucoma prevention and treatment. Given the favorable side-effect profile and extensive safety data regarding this class of medication, our results provide a preliminary impetus for providers to consider GLP-1R agonists as preferred therapy for patients with DM and at high risk for glaucoma.

## Data Availability

All data are available upon reasonable request.

---

**Supplemental Table 1.**
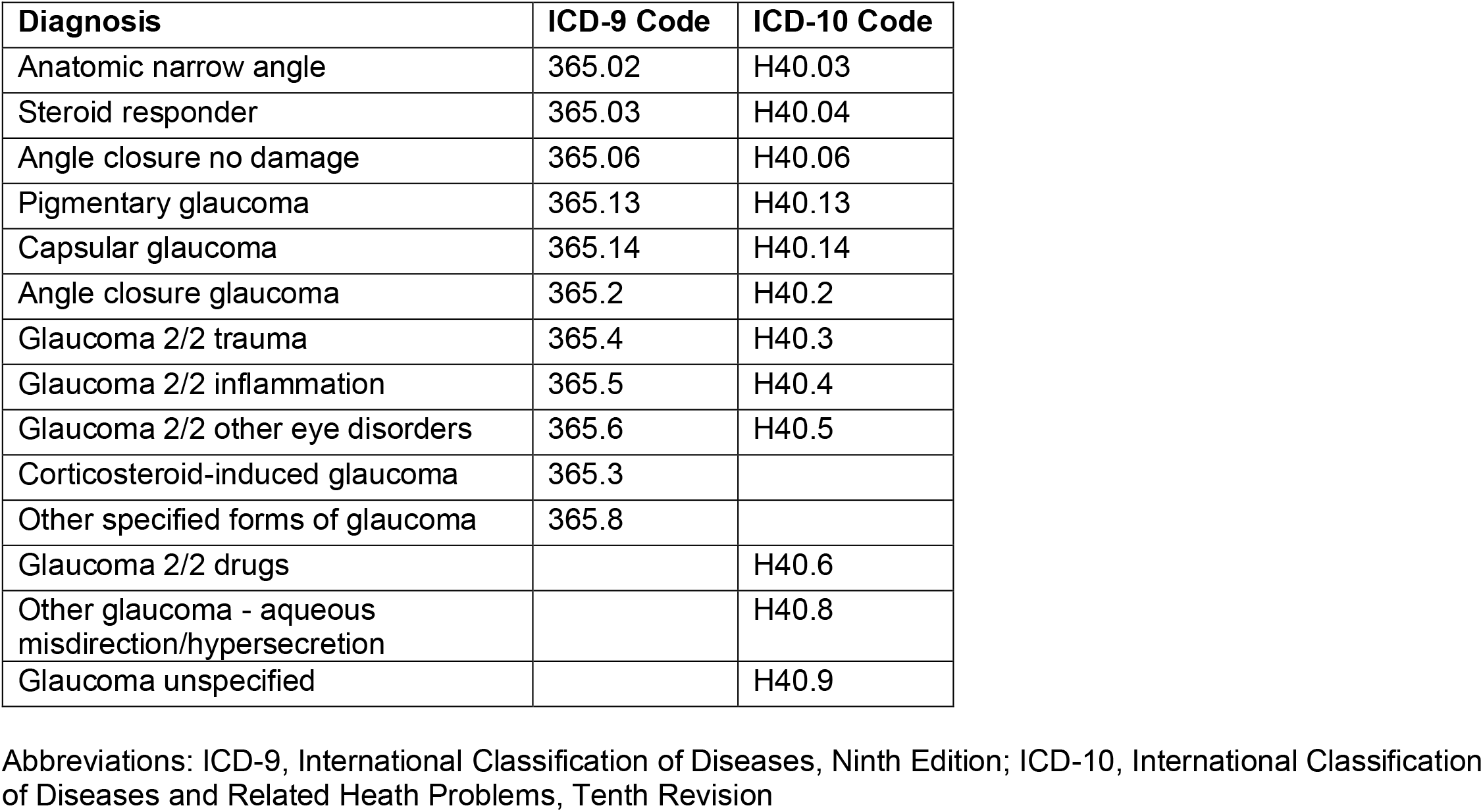
ICD-9/ICD-10/CPT codes used in the regular analysis.

